# Pure-tone audiometry and dichotic listening in primary progressive aphasia and Alzheimer’s disease

**DOI:** 10.1101/2024.03.14.24304280

**Authors:** Jessica Jiang, Jeremy Johnson, Benjamin A Levett, Lucy B Core, Anna Volkmer, Nehzat Koohi, Doris-Eva Bamiou, Charles R Marshall, Jason D Warren, Chris JD Hardy

**Author notes:** These authors contributed equally to the work (Jessica Jiang, Jeremy Johnson). These authors contributed equally to the work (Jason D Warren, Chris JD Hardy).

## Abstract

Hearing is multifaceted and the relative contributions of peripheral and central hearing loss are rarely considered together in the context of dementia. Here, we assessed peripheral (as measured with pure-tone audiometry) and central (as measured with dichotic listening) hearing in 19 patients with typical amnestic Alzheimer’s disease (tAD), 10 patients with logopenic variant primary progressive aphasia (lvPPA), 11 patients with nonfluent/agrammatic variant PPA (nfvPPA), 15 patients with semantic variant PPA (svPPA), and 28 healthy age-matched individuals. Participants also underwent neuropsychological assessment and magnetic resonance image scanning, allowing us to use voxel-based morphometry to assess associations between hearing scores and grey matter volume. Dichotic listening was impaired in all patient groups relative to healthy controls. In the combined patient (but not healthy control) cohort, dichotic listening scores were significantly correlated with measures of global cognitive functioning and speech-based neuropsychological tasks. Pure-tone audiometry scores were not significantly elevated in any patient group relative to the healthy control group, and no significant correlations were observed between peripheral hearing and neuropsychological task performance in either the combined patient or healthy control cohorts. Neuroanatomically, dichotic listening performance was associated with grey matter volume in a bilateral fronto-temporo-parietal network over the combined patient cohort, but no correlates were identified for pure-tone audiometry. Our findings highlight the importance of speech parsing mechanisms beyond elementary sound detection in driving cognitive test performance, underline the importance of assessing central hearing alongside peripheral hearing in people with dementia, and further delineate the complex auditory profiles of neurodegenerative dementias.

## Introduction

There is now a plethora of evidence suggesting that hearing loss and dementia are linked. Peripheral hearing impairment measured with pure-tone audiometry has been identified as a major risk factor for dementia in mid-life (Lin et al., 2011; Livingston et al., 2020) and recent evidence suggests that peripheral amplification may improve quality of life and hearing abilities in people with dementia (Leroi et al., 2020), and may alter cognitive trajectories in older adults at increased risk of cognitive decline (Bucholc et al., 2021, 2022; Lin et al., 2023; Maharani et al., 2018; Yeo et al., 2023). Several potential mechanisms have been proposed that may account for this association (Griffiths et al., 2020; Johnson, Marshall, et al., 2020). One important observation is that peripheral hearing loss is typically associated with poorer cognitive performance in healthy older individuals, participants with mild cognitive impairment and established dementia (Golub et al., 2020; Loughrey et al., 2018; Taljaard et al., 2015). Prevailing evidence has suggested that these associated cognitive deficits are not purely attributable to the inability to hear test instructions or stimuli, as impairments have been identified for tests that both do and do not require speech perception for accurate performance. In a large meta-analysis and systematic review of forty studies from twelve countries, Loughrey and colleagues (2018) identified a small but significant association between age-related hearing loss (as measured with pure-tone audiometry) and all cognitive function domains included in the analyses.

However, other findings have been more equivocal. A recent study of 55 people with established Alzheimer’s disease (AD) found that pure-tone audiometry was not significantly associated with performance on the preclinical Alzheimer’s cognitive composite (PACC) (Donohue et al., 2014), which combines tests assessing episodic memory, executive function and global cognition, all of which are neuropsychological domains that are affected early in the course of AD (Martínez-Dubarbie et al., 2023). A study of 368 cognitively healthy individuals aged approximately 70 years found that pure-tone audiometry performance only predicted Mini-Mental State Examination (MMSE) score when an item requiring repetition of a single phrase was included; and there were no significant correlations between pure-tone audiometry and any other cognitive measures (Parker et al., 2019).

Furthermore, peripheral hearing impairment is only part of the picture: hearing loss attributable to involvement of central auditory pathways (i.e., beyond what can be explained by pure-tone audiometry performance) has been noted in different forms of dementia, notably AD and primary progressive aphasia (PPA). Such central hearing impairment has been demonstrated by tests designed to measure cortical auditory processes (such as the Queen Square Tests of Auditory Cognition) (Gates et al., 2015; Grube et al., 2016; Hardy et al., 2020; Jiang et al., 2023; Johnson, Marshall, et al., 2020). These findings highlight reverse causation as a possible mechanism for this complex hearing loss, i.e. that early brain changes associated with neurodegenerative dementias affect parts of the auditory brain network, which manifest as symptoms of hearing loss that are undetectable with standard audiometric testing (Cope et al., 2015; Musiek et al., 2007, 2017).

Dichotic listening tasks have been widely used to probe central hearing function, and patients with AD are consistently impaired relative to healthy older listeners on these tasks (Bouma & Gootjes, 2011; Häggström et al., 2018, 2020; Idrizbegovic et al., 2011, 2013; Utoomprurkporn et al., 2020), even when peripheral hearing function is controlled for (Gates et al., 2010; Mohammed et al., 2022). Dichotic listening involves the simultaneous presentation of different acoustic events to each ear (Broadbent, 1954; Cherry, 1953). Several clinical tests have been developed, such as the widely known dichotic digits test (Musiek, 1983; Musiek et al., 1991), in which pairs of digits are presented dichotically, with one digit from each pair presented to the left ear and a different digit presented to the right ear, simultaneously.

Previous research has also consistently shown that participants are more accurate in identifying verbal stimuli that are presented to the right ear in the context of dichotic listening tasks (Kimura, 1961). This benefit is termed the ‘right-ear advantage’ and reflects preferential processing of verbal information from the right ear by the dominant left cerebral hemisphere. Intriguingly, the right-ear advantage has been found to be amplified in patients with dementia compared with age-matched healthy individuals (Duchek & Balota, 2005; Idrizbegovic et al., 2011; Strouse et al., 1995). In a recent meta-analysis, participants living with all-cause dementia had a dichotic digit mean score in the right ear approximately 20% higher than in the left ear (Utoomprurkporn et al., 2020).

Although pure-tone audiometry and dichotic listening tests perhaps represent the most widely used clinical tests of peripheral and central hearing function, respectively, to our knowledge no studies have directly compared performance on these tasks in well-characterised dementia cohorts. In particular, dichotic listening performance has never been reported in PPA, the rare group of language-led dementias. This group is of considerable interest here given that PPA predominantly affects the left hemisphere in its earliest stages (Gorno-Tempini et al., 2011; Lombardi et al., 2021; Marshall et al., 2018) and has been shown in previous research to have a diverse profile of cortical auditory impairments (Bozeat et al., 2000; Goll et al., 2010; Grube et al., 2016; Hardy et al., 2017, 2018, 2019; Jiang et al., 2022, 2023; Johnson, Jiang, et al., 2020). However, abnormal pure-tone audiometry profiles have previously been identified in patients with nonfluent/agrammatic variant PPA (nfvPPA) (Hardy et al., 2019), implying a complex interaction between ‘peripheral’ and ‘central’ hearing processes (e.g., efferent auditory pathway dysfunction) in this form of cortical degeneration.

Here, we aimed to measure peripheral hearing (as measured with pure-tone audiometry) alongside central hearing (as measured with dichotic listening) in patients with typical amnestic Alzheimer’s disease and all major PPA syndromes in comparison with healthy older individuals. We hypothesised that patients with nfvPPA would have elevated pure-tone audiometry thresholds in comparison with healthy individuals and other patient groups, based on our previous findings (Hardy et al., 2019); and that all patient groups with the exception of semantic variant (sv)PPA (considering brain regions chiefly implicated in dichotic listening are relatively spared) would perform significantly worse than healthy individuals on the dichotic listening task. We further hypothesised that patient groups with left-lateralised atrophy affecting temporoparietal cortex (i.e., nfvPPA and logopenic variant (lv)PPA) would show an attenuated dichotic listening right-ear advantage compared to healthy individuals.

We also assessed how performance on both hearing tasks related to performance on a battery of standard neuropsychological tests. Cognitive processes that depend on active parsing of incoming speech signals (e.g., to enter auditory working memory) are likely to be affected by impairments of auditory parsing mechanisms (as indexed by dichotic listening). Speech audibility (i.e., the ability to detect speech signals) is required for successful cognitive processing but alone does not guarantee successful parsing of speech signals (Holmes & Griffiths, 2019), leading us to hypothesise that dichotic digits test performance would correlate with speech-dependent cognitive test performance while pure-tone audiometry would not.

Additionally, we assessed the structural neuroanatomical associations of hearing test performance (peripheral and central) in the combined AD and PPA cohort, using voxel-based morphometry on patients’ brain magnetic resonance imaging (MRI) scans. Based on previous research findings, we hypothesised that pure-tone audiometry might be correlated with regional grey matter atrophy in cortical areas (primary auditory cortex, medial temporal lobe and posterior superior temporal regions) showing atrophy due to prolonged auditory deafferentation (Armstrong et al., 2019; Eckert et al., 2012; Lin et al., 2014; Parker et al., 2019; Ren et al., 2018; Xu et al., 2019), while dichotic listening would be correlated with regional grey matter atrophy in cortical regions (posterior superior temporal, parietal and prefrontal cortices) where disease-related atrophy produces a (central) hearing deficit (Hardy et al., 2018; Hugdahl & Westerhausen, 2016; Jäncke et al., 2001; Jiang et al., 2023; Johnson, Jiang, et al., 2020; Thomsen et al., 2004).

## Methods

### Participants

Nineteen patients with typical amnestic Alzheimer’s disease (tAD), 10 patients with logopenic variant primary progressive aphasia (lvPPA), 11 patients with nonfluent/agrammatic variant primary progressive aphasia (nfvPPA), and 15 patients with semantic variant primary progressive aphasia (svPPA) were recruited via a specialist cognitive clinic. All patients fulfilled consensus clinical diagnostic criteria (Dubois et al., 2014; Gorno-Tempini et al., 2011) with clinically mild-to-moderate disease, supported by MRI showing compatible brain atrophy profiles without significant cerebrovascular burden. Twenty-nine healthy age-matched individuals with no history of neurological or psychiatric disorders were recruited from the Dementia Research Centre volunteer database. No participant had a history of otological disease, other than presbycusis.

All participants gave informed consent to take part in the study. Ethical approval was granted by the UCL-NHNN Joint Research Ethics Committee (Approval ID 06NO32), in accordance with Declaration of Helsinki guidelines.

### Peripheral hearing assessment

Following British Society of Audiology guidelines (BSA, 2018), pure-tone audiometry was performed at 250, 500, 1000, 2000, 4000 and 8000 Hz using either a dual-channel GSI Audiostar Pro audiometer or an Amplivox Screening audiometer model 116, with calibrated, noise-reducing headphones in a quiet room. The pure-tone threshold for each ear was calculated as the average minimum threshold in decibels hearing level (dB HL) across the 500, 1000, 2000 and 4000 Hz frequencies most relevant to speech processing (Lin & Reed, 2021).

### Dichotic listening task

Dichotic listening was assessed using the dichotic digits test (Musiek, 1983), administered from a Dell Latitude laptop computer running Experiment Builder software (*SR Research Experiment Builder 2.3. 1 [Computer Software].*, 2020) and using Audio-Technica M50x headphones in a quiet room. Prior to commencing the task, participants were played a 1kHz test tone, asked to confirm that they could hear this tone in both ears, and invited to adjust the volume to a comfortable listening level (at least 70dB). On each trial, two pairs of different digits were presented to the participant; the digits in each pair overlapped where one digit was presented to the left ear and the other to the right ear. After each trial, the participant was asked to repeat the digits that they heard, in any order. Twenty trials were administered, with a maximum score of four on each trial (one per digit), yielding a maximum possible score of 80.

### Neuropsychological assessment

A general neuropsychological battery was administered to participants alongside their hearing assessments. The researcher ensured that all spoken instructions and practice trials were delivered at a sound level that was easily audible for each participant, and participants were allowed to wear hearing aids if preferred.

Tests that did not require speech perception comprised:

1. *Executive function:* The Wechsler Adult Scale of Intelligence (WASI) Matrix Reasoning test (Wechsler, 1999). For each item, the participant is presented with a panel typically representing a series of figures at the top of the page in which there is a pattern, with one figure in the series left incomplete. The participant has to select which figure would complete the pattern correctly from an array of five possible options.
2. *Executive function:* Verbal and category fluency tests (Delis et al., 2001). For the verbal fluency task, participants are given sixty seconds to name as many words they can beginning with the letter “F”. Proper nouns are not permitted. For the category fluency task, participants are given sixty seconds to name as many words belonging to the category of “animal”.
3. *Semantic memory:* The British Picture Vocabulary Scale (BPVS) (Dunn, L & Whetton, 1982). This is a word-picture matching task where a participant is shown a word that is also read aloud by the experimenter. They are asked to select the picture that best matches the meaning of the word from four possible options.
4. *Episodic memory:* The Camden Paired Associates Learning (PAL) test (Warrington, 1996). Participants are confronted with written word pairs in three sets of eight, which the experimenter reads out loud and the participant repeats. Each word pairing is shown for three seconds. After the presentation of the set of eight pairs of words, the participants are immediately (∼1 minute delay to test) presented with the first word from each pair and asked to name the word that was paired with it.
5. *Visual Perception:* The Object Decision test from the Visual Object and Space Perception (VOSP-OD) battery (Warrington & James, 1991). Participants are shown four silhouette drawings and asked to indicate which of the drawings represents a “real” object.

Tests relying on speech perception for successful completion comprised:

1. *Arithmetic*: The Graded Difficulty Arithmetic (GDA) test (Jackson & Warrington, 1986). The experimenter verbally poses a series of arithmetical calculations which the participant must answer verbally. There are twelve items involving addition and twelve items involving subtraction with a 10-second time limit for each item.
2. *Auditory verbal working memory:* The forward and reverse digit span tests (Wechsler, 1987). These require the participant to repeat a string of digits of increasing length spoken by the experimenter in the same (forward condition) or reverse (backwards condition) order.

### Analysis of clinical and neuropsychological data

Data were analysed in Stata v14. For continuous demographic and neuropsychological data, participant groups were compared using analysis of variance (ANOVA) tests; categorical data were compared using Fisher’s exact tests. Where the initial omnibus test showed a significant effect of diagnosis, post-hoc t-tests were conducted to identify the groups driving the effect; where the initial omnibus test was not significant, no post-hoc tests were conducted.

### Analysis of peripheral and dichotic listening data

For peripheral hearing, each individual participant’s ‘better ear average’ was calculated by averaging across the 500-4000Hz frequencies of each ear and taking the lower and higher values, respectively. This 4-frequency average in the better ear was adopted as the main measure of peripheral hearing, following previous recommendations (Lin & Reed, 2021), and all subsequent correlations and additional analyses incorporating pure-tone audiometry performance are based on this measure unless otherwise specified. However, we also calculated an average across the full range of frequencies assessed (250Hz-8000Hz) and a high-frequency average across the frequencies 4000Hz and 8000Hz (see Table S1). Better-hearing ear lateralisation was analysed as a categorical variable.

For dichotic listening, we obtained a total score on the dichotic digits test, as well as considering a right-ear advantage score. This was calculated by subtracting the number of correctly repeated digits presented to the right ear minus the number of correctly repeated digits presented to the left ear.

Participant groups were compared on peripheral and dichotic listening using analysis of covariance (ANCOVA) models. Analyses using a peripheral hearing score as a dependent variable were adjusted for covariates of age, sex and forward digit span forward (as a proxy of both auditory verbal working memory and overall severity of cognitive impairment across groups); analyses using dichotic listening score as a dependent variable were additionally adjusted for better ear average. In parallel analyses we ran the same models without including forward digit span score as a covariate (see Table S2).

We analysed right-ear advantage scores at two levels. First, we conducted within-subjects one-tailed one-sample t-tests to assess whether the right-ear advantage was significantly different from 0 (i.e. indicating the presence of a right-ear advantage if positive) in each participant group. Second, we conducted an ANCOVA model, as described above, to assess whether there was a between-subjects effect of diagnosis on right-ear advantage. Categorical data were compared using unadjusted Fisher’s exact tests.

### Associations between peripheral hearing, dichotic listening and neuropsychological variables

Pearson correlation coefficients were calculated to assess the relationship between peripheral hearing (better ear average) and dichotic listening, and the associations between both hearing tests and MMSE score, in the healthy individuals and combined patient cohort separately. Coefficients were also calculated between peripheral hearing and each neuropsychological test as well as between dichotic listening and each neuropsychological test. We additionally ran partial correlation analyses adjusting for better ear average for any significant correlations between dichotic listening and neuropsychological task performance to ensure that these were not accounted for by peripheral hearing.

Here we assessed significance using a Bonferroni correction for multiple comparisons such that the 0.05 threshold for significance was divided by nine (the number of individual neuropsychological tasks; Table 1; Figure 2; Figure S2) to give an adjusted threshold of p < 0.0056.

**Table 1.**
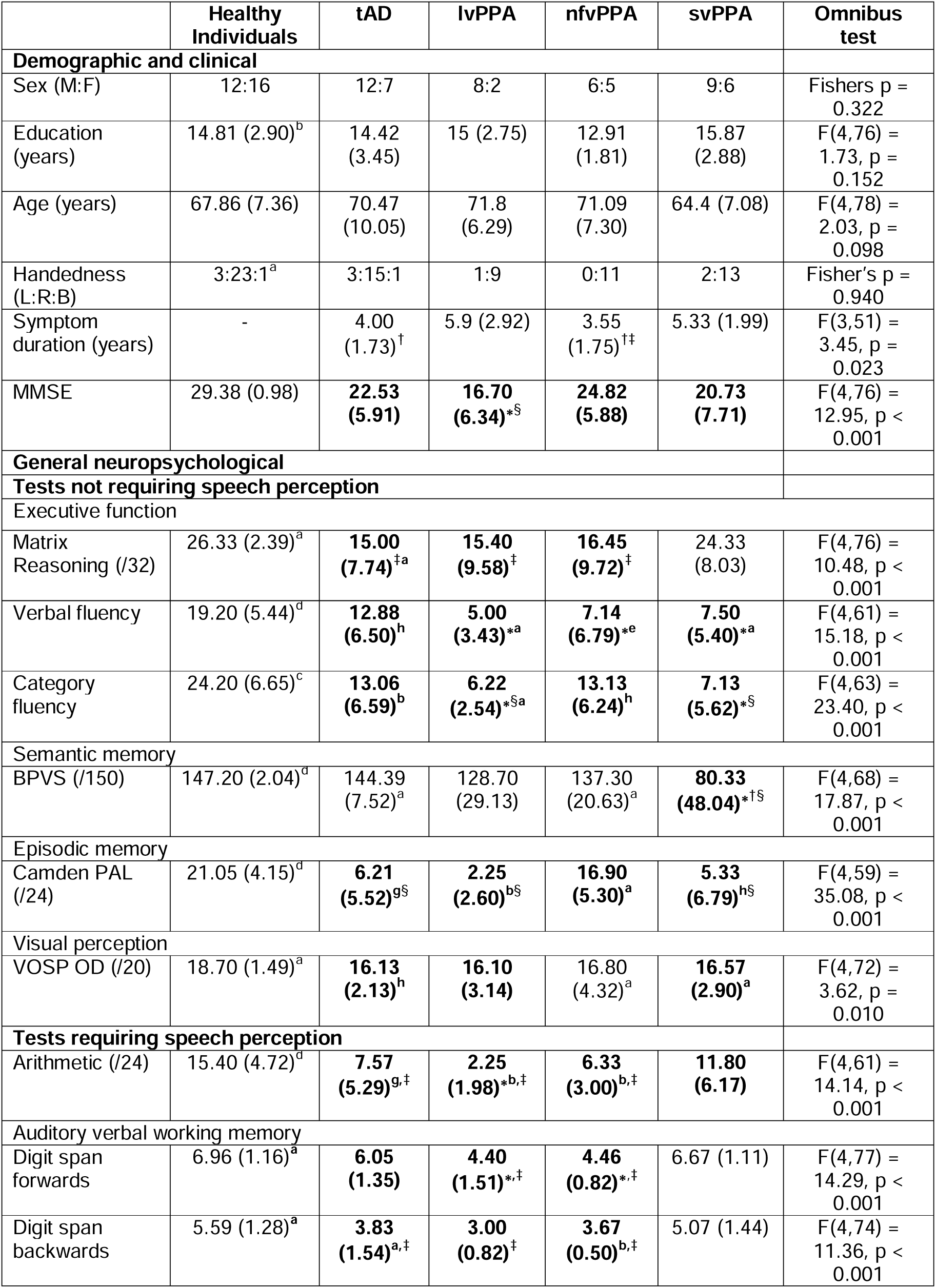
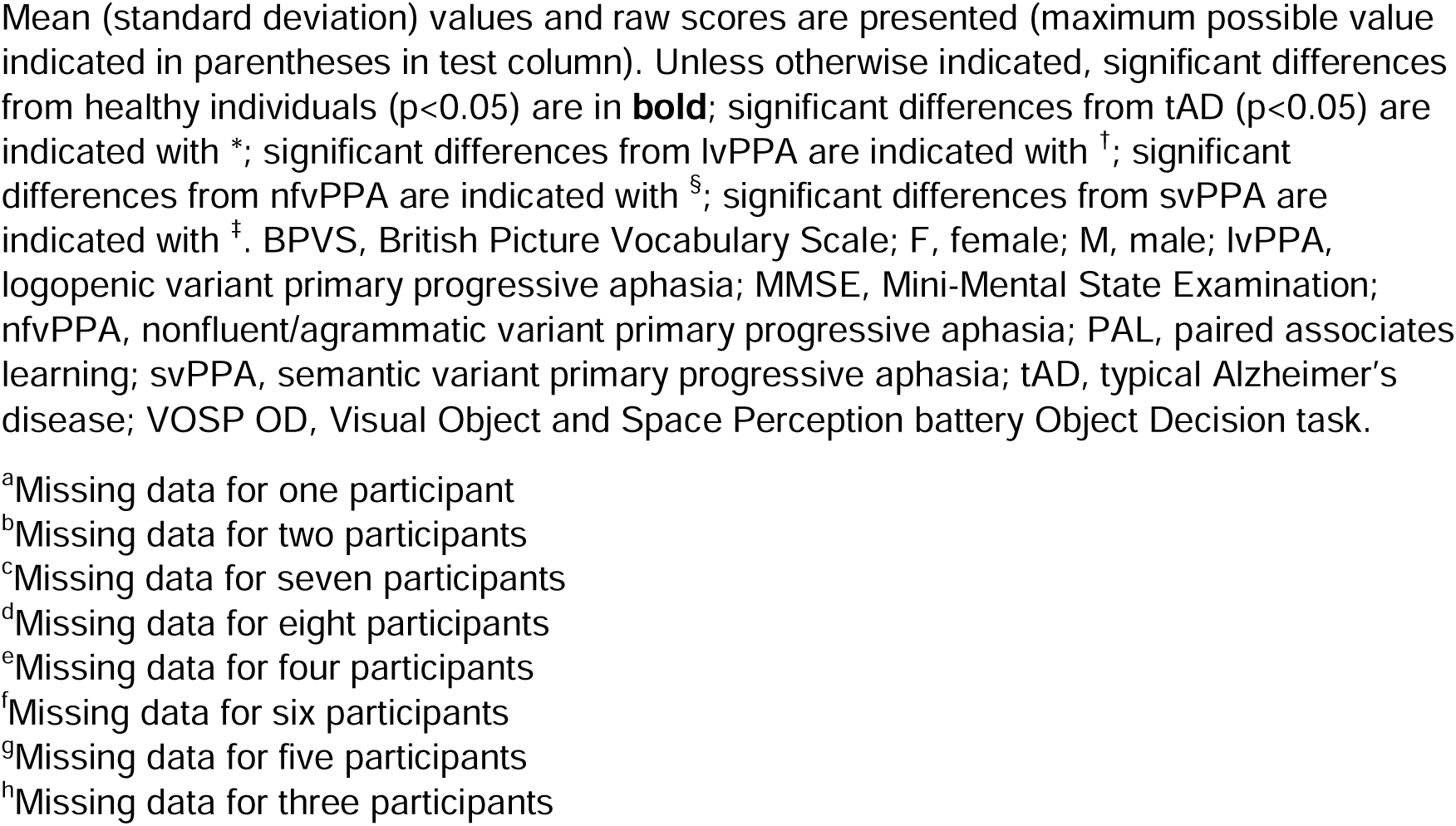
General demographic, clinical and neuropsychological characteristics of participant groups.

### Brain image acquisition and analysis

Volumetric brain magnetic resonance images were acquired for 45 patients (15 AD, 7 lvPPA, 9 nfvPPA, 14 svPPA) in a 3T Siemens Prisma MRI scanner, using a 64-channel phased array head coil and following a T1-weighted sagittal 3D magnetization prepared rapid gradient echo (MPRAGE) sequence (echo time = 2.9 ms, inversion time = 900 ms, repetition time = 2200 ms), with dimensions 256 mm × 256 mm × 208 mm and voxel size 1.1 mm × 1.1 mm × 1.1 mm.

For the voxel-based morphometry (VBM) analysis, patients’ brain images were first pre-processed and normalised to MNI space using Statistical and Parametric Mapping (SPM) software v12 and the DARTEL toolbox with default parameters running under MATLAB R2014b. Images were smoothed using a 6mm full width at half-maximum (FWHM) Gaussian kernel. Total intracranial volume was calculated for each patient by summing white matter, grey matter and CSF volumes post-segmentation (Malone et al., 2015), in order to control for individual differences in pre-morbid brain size. We used an automatic mask-creating strategy to create an explicit, study-specific brain mask (Ridgway et al., 2009). A study-specific mean brain template image was created by warping all patients’ native-space whole-brain images to the final DARTEL template and using the ImCalc function to generate an average of these images. During preprocessing, significant movement artefacts were identified in the scan for one tAD participant, meaning that 44 scans were ultimately included in these analyses.

We assessed grey matter associations of peripheral hearing (better ear average) and dichotic listening (total performance and right-ear advantage score) over the combined patient cohort. Voxel-wise grey matter intensity was modelled as a function of hearing score in a multiple regression design, incorporating covariates of diagnostic group membership, age, total intracranial volume, MMSE score and sex for the peripheral hearing analyses as well as the additional covariate of better ear average for the dichotic listening analyses. Negative contrasts were assessed for the peripheral hearing analyses (as higher scores here indicate worse hearing), and positive contrasts for the dichotic listening analyses. First, statistical parametric maps were generated at P < 0.05_FWE_ threshold over the whole brain. Additionally, statistical parametric maps were generated using an initial cluster-defining threshold (p < 0.001) and assessed at peak-level significance threshold p<0.05 after family-wise error (FWE) correction for multiple voxel-wise comparisons within six separate predefined regions of interest, based on previously published work on hearing in the healthy brain and in neurodegenerative disease (Eckert et al., 2012; Griffiths et al., 2020; Hardy et al., 2018; Hugdahl & Westerhausen, 2016; Jäncke et al., 2001; Jiang et al., 2023; Johnson, Jiang, et al., 2020; Ren et al., 2018; Thomsen et al., 2004). These regions comprised (i) Heschl’s gyrus, (ii) the posterior superior temporal gyrus and planum temporale; (iii) medial temporal lobe (whilst not a canonical auditory area, previous work has suggested that atrophy here may correlate with reduced pure-tone audiometry performance); (iv) angular gyrus and supramarginal gyrus; (v) superior parietal lobe; and (vi) prefrontal cortex (superior frontal gyrus, middle frontal gyrus, inferior frontal gyrus). Due to the asymmetry of these disease groups, regions in the left and right cerebral hemispheres were analysed separately. Anatomical volumes were derived from Oxford-Harvard cortical maps (Desikan et al., 2006) and are shown in Supplementary Figure S1.

## Results

General participant and neuropsychological performance data by group are presented in Table 1. Peripheral and dichotic listening characteristics by group are presented in Table 2 and Figure 1. Pearson correlation values between hearing and cognitive scores are presented in Figure 2 and Figure S2.

**Table 2.**
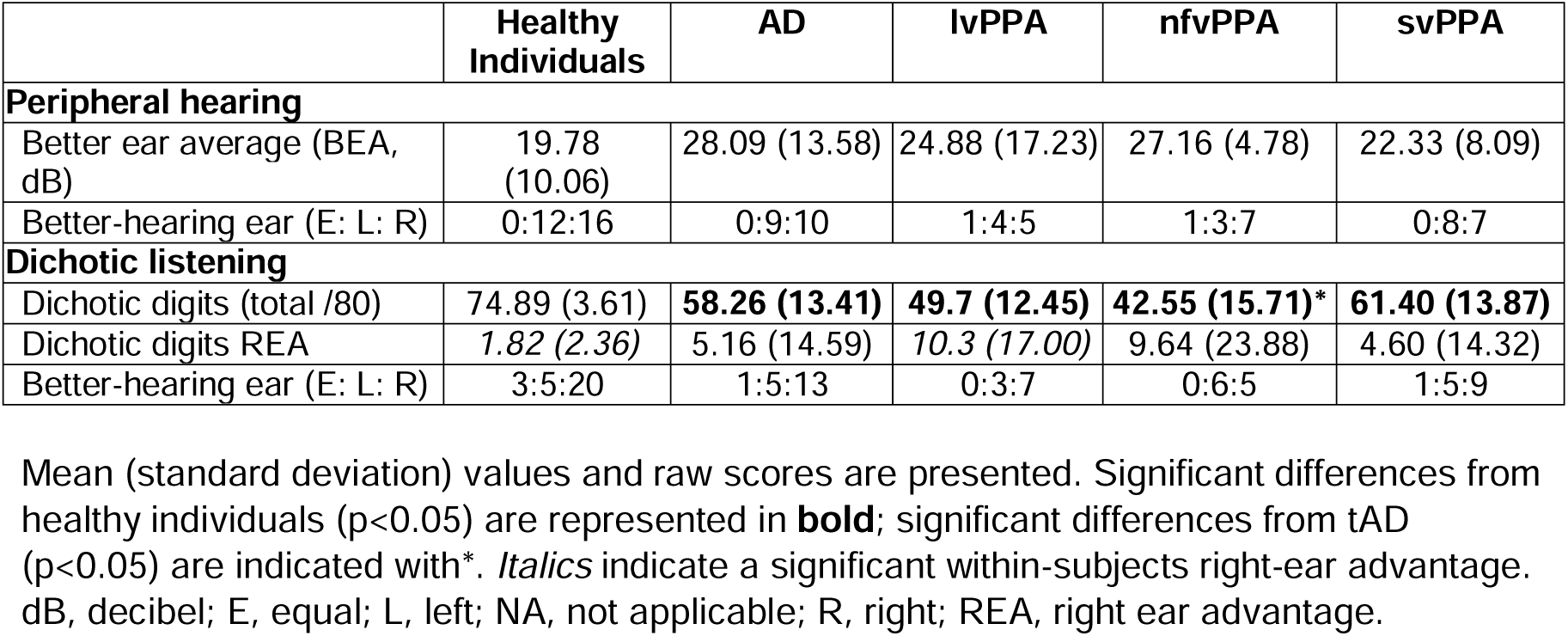
Peripheral and dichotic listening characteristics of participant groups.

|  | Healthy Individuals | AD | IvPPA | nfvPPA | svPPA |
| --- | --- | --- | --- | --- | --- |
| **Peripheral hearing** |  |  |  |  |  |
| Better ear average (BEA, dB) | 19.78 (10.06) | 28.09 (13.58) | 24.88 (17.23) | 27.16 (4.78) | 22.33 (8.09) |
| Better-hearing ear (E: L: R) | 0:12:16 | 0:9:10 | 1:4:5 | 1:3:7 | 0:8:7 |
| **Dichotic listening** |  |  |  |  |  |
| Dichotic digits (total /80) | 74.89 (3.61) | **58.26 (13.41)** | **49.7 (12.45)** | **42.55 (15.71)\*** | **61.40 (13.87)** |
| Dichotic digits REA | 1.82 (2.36) | 5.16 (14.59) | 10.3 (17.00) | 9.64 (23.88) | 4.60 (14.32) |
| Better-hearing ear (E: L: R) | 3:5:20 | 1:5:13 | 0:3:7 | 0:6:5 | 1:5:9 |

Mean (standard deviation) values and raw scores are presented. Significant differences from healthy individuals ( $p < 0.05$ ) are represented in **bold**; significant differences from tAD ( $p < 0.05$ ) are indicated with\*. *Italics* indicate a significant within-subjects right-ear advantage. dB, decibel; E, equal; L, left; NA, not applicable; R, right; REA, right ear advantage.

**Figure 1.**
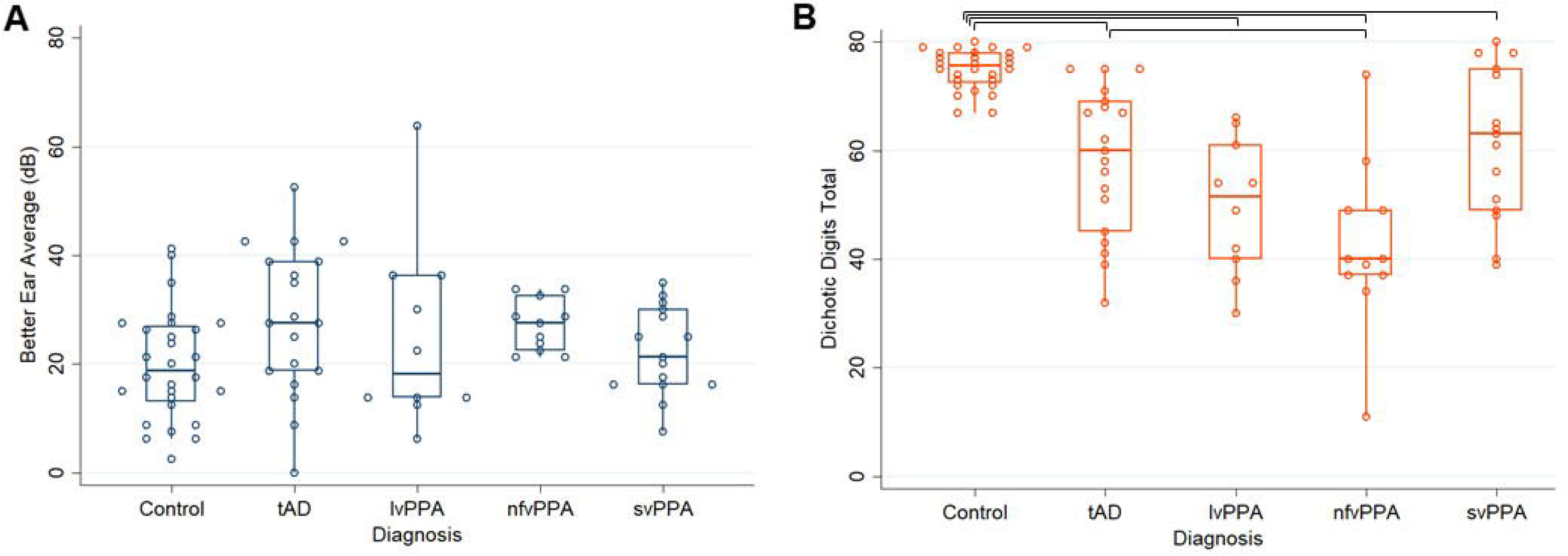
Box-and-whisker plots showing **A)** better ear average measured by pure-tone audiometry (i.e., higher values indicate poorer hearing) for all participant groups; and **B)** dichotic listening scores for all participant groups. Boxes code interquartile range; whiskers code 95% confidence intervals; transverse lines code median scores; dots show individual scores. Significant between-group differences (after adjustment for covariates; see text) are represented using horizontal brackets above the relevant groups. Control, healthy individual cohort; lvPPA, logopenic variant primary progressive aphasia; nfvPPA, nonfluent/agrammatic variant primary progressive aphasia; svPPA, semantic variant primary progressive aphasia; tAD, typical Alzheimer’s disease.

**Figure 2.**
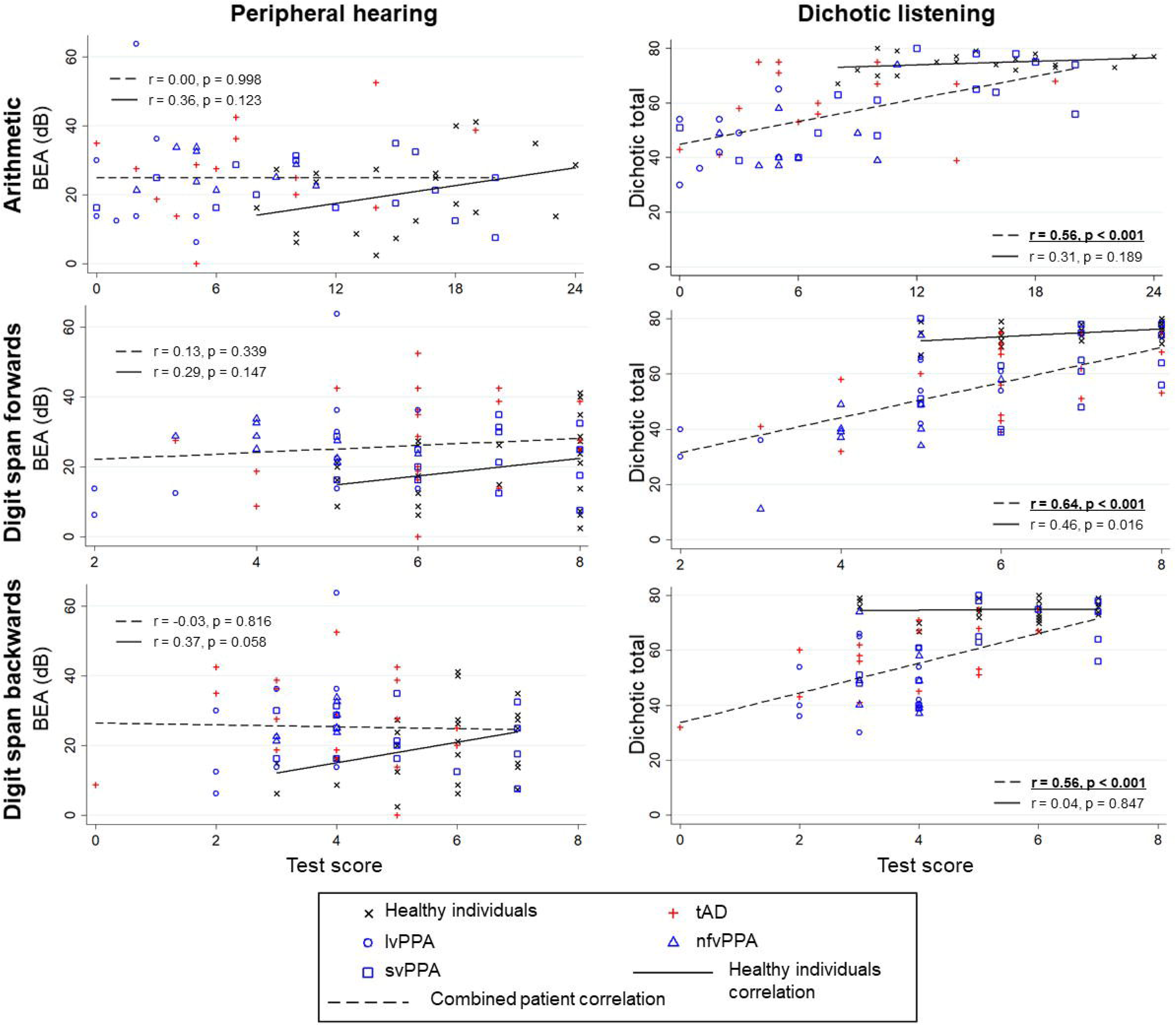
Scatter plots showing correlations between peripheral hearing ((better ear average); left panels) and dichotic listening (total score; right panels) with neuropsychological tests requiring speech perception for successful performance in patient groups and healthy individuals. Diagnostic group membership is described in the key. For peripheral hearing, a lower pure tone audiometric average indicates better hearing so here negative *r* values would indicate that better peripheral hearing performance is associated with better psychometric performance. For central hearing, a higher dichotic digit total score indicates better hearing so here positive *r* values would indicate that better central hearing performance is associated with better psychometric performance. Significant correlations (after correction for multiple comparison; see Methods) are indicated in **<u>bold underline</u>**. Correlations for tests not requiring speech perception for successful performance are displayed in Supplementary Figure S2. BEA, better ear average; control; healthy individual cohort; tAD, typical Alzheimer’s disease; lvPPA, logopenic variant primary progressive aphasia; nfvPPA, nonfluent/agrammatic variant primary progressive aphasia; svPPA, semantic variant primary progressive aphasia.

One healthy control participant was identified as an outlier on the dichotic listening test (scoring more than five standard deviations lower than the second-lowest performing healthy individual) and so was removed from all subsequent analyses, leaving 28 healthy individuals.

### General participant group characteristics

Participant groups did not differ significantly in sex distribution, age, handedness or years of education (all p > 0.05; Table 1). Patient groups did differ in symptom duration, with lvPPA and svPPA groups having a significantly longer symptom duration than the nfvPPA group, and the lvPPA group having a significantly longer symptom duration than the tAD group. Groups also differed significantly in terms of MMSE score, driven by healthy individuals scoring significantly better than each patient group and the lvPPA group performing significantly worse than the tAD and nfvPPA group (Table 1).

### Neuropsychological performance across participant groups

#### Tests not requiring speech perception

Participant groups differed significantly on WASI Matrix reasoning scores (F(4,76) = 10.48, p < 0.001), driven by the healthy individuals and svPPA groups performing better than each other patient group (all p < 0.05). There was a main effect of diagnosis on both fluency tests (verbal fluency, F(4,61) = 15.18; category fluency, F(4,63) = 23.40; both p < 0.001) accounted for by the healthy individuals on average scoring significantly higher than all other participant groups on both tasks (all p < 0.001) (Table 1). The omnibus test for BPVS was significant (F(4,68) = 17.87, p < 0.001), and here the svPPA group scored significantly lower on average than every other group (all p < 0.001). Camden PAL scores also differed significantly across participant groups (F(4,59) = 35.08, p < 0.001), with each patient group scoring significantly worse than healthy individuals (all p < 0.05), and the nfvPPA group scoring significantly higher than each of the other patient groups (all p < 0.05). There was also a main effect of diagnosis on VOSP OD scores (F(4,72) = 3.62, p = 0.010), with the tAD, lvPPA and svPPA groups scoring significantly worse than the healthy individuals (all p < 0.05).

#### Tests requiring speech perception

Participant groups differed significantly on arithmetic (F(4,61) = 14.14, p < 0.001), driven by the healthy individuals having greater scores than each patient group; and the svPPA group scoring better than the other patient groups (all p < 0.05). Groups also differed significantly on forward digit span (F(4,77) = 14.29, p < 0.001); all patient groups except svPPA scored significantly worse than healthy individuals (all p < 0.05); and the lvPPA and nfvPPA groups scoring significantly lower than the tAD and svPPA patient groups (all p < 0.05). The effect of diagnosis on reverse digit span was also significant (F(4,74) = 11.36, p < 0.001), with the tAD, lvPPA and nfvPPA groups all scoring significantly worse than the healthy individuals and svPPA groups (all p < 0.05).

### Peripheral and dichotic listening characteristics of participant groups

For better ear average, the overall model adjusting for age, sex and digit span forward score was significant (F(7,74) = 5.07, p < 0.001), however whilst age was significantly associated with better ear average (t = 3.73, p < 0.001), digit span forward score, sex and diagnosis (all p>0.05) were not (Supplementary Table S2). Dropping forward digit span as a covariate did not substantially change these results (Supplementary Table S2). Parallel analyses for better ear averages over different frequency ranges (250Hz – 8000Hz and 4000Hz – 8000Hz yielded qualitatively similar results (Supplementary Table S1). Participant groups also did not differ significantly in terms of better-hearing ear lateralisation (Table S2).

For dichotic listening score, the overall model adjusting for age, sex, digit span forward score and better ear average was significant (F(8,73) = 18.49, p < 0.001). Sex (t = 2.65, p = 0.010), better ear average (t=2.63, p=0.010), digit span forward score (t = 5.16, p < 0.001) and diagnosis (t = 2.23, p = 0.001) were significantly associated with dichotic listening performance whilst age (p>0.05) was not (Table S2). Post-hoc tests showed that the tAD (t=-2.71, p=0.008), lvPPA (t=-2.00, p=0.049), nfvPPA (t=-3.94, p<0.001), and svPPA (t=-3.04, p=0.003) groups all performed worse than healthy individuals (Table 2), with the nfvPPA group also performing significantly worse than the tAD group (t=-2.18, p=0.033). Dropping forward digit span as a covariate did not substantially change these results (Supplementary Table S2).

One-sample t-tests indicated a significant right-ear advantage in healthy individuals (t(27) = 4.09, p < 0.001) and the lvPPA group (t(9) = 1.92, p = 0.044), but not in the other participant groups (all p > 0.05) (Table 2). An overall model for right-ear advantage score comparing across diagnostic groups adjusting for age, sex, digit span forward score and better ear average was not significant (Table S2). The proportion of participants who showed a right ear advantage also did not differ significantly across groups (Table S2).

### Associations between peripheral hearing, dichotic listening and neuropsychological variables

#### Peripheral hearing and dichotic listening

Peripheral hearing ability (better ear average) was not significantly associated with dichotic listening performance in the healthy individual group (*r* = -0.18, p = 0.350), nor in the combined patient cohort (*r* = -0.08, p = 0.588).

#### Peripheral hearing and MMSE

MMSE score was not significantly associated with peripheral hearing ability (better ear average) in the healthy individual (*r* = 0.21, p = 0.305) or combined patient cohort (*r* = 0.02, p = 0.878).

#### Dichotic listening and MMSE

MMSE score was not significantly associated with dichotic listening score in the healthy individual group (*r* = -0.23, p = 0.267). There was a significant association in the combined patient cohort (*r =* 0.40, p = 0.003).

#### Peripheral hearing and neuropsychological variables

Figure 2 and Figure S2 show the Pearson correlation coefficients for associations between peripheral and neuropsychological test performance. No significant associations were observed for either the healthy individuals or combined patient cohorts (all p > 0.0056).

#### Dichotic listening and neuropsychological variables

Figure 2 and Figure S2 also show the Pearson correlation coefficients for associations between dichotic listening total score and neuropsychological test performance. No significant associations were seen in the healthy individual group (all p > 0.0056). In the combined patient group, all three of the tasks requiring speech perception for successful completion were significantly associated with dichotic listening performance after multiple comparison correction (Figure 2). These correlations remained significant when controlling for peripheral hearing ability (see Supplementary Materials).

### Neuroanatomical data

Statistical parametric maps of grey matter regions associated with peripheral hearing and dichotic listening are shown in Figure 3 and local maxima are summarized in Table. Across the combined patient cohort, pure-tone audiometry (better ear average) was not significantly associated with regional grey matter volume.

**Figure 3.**
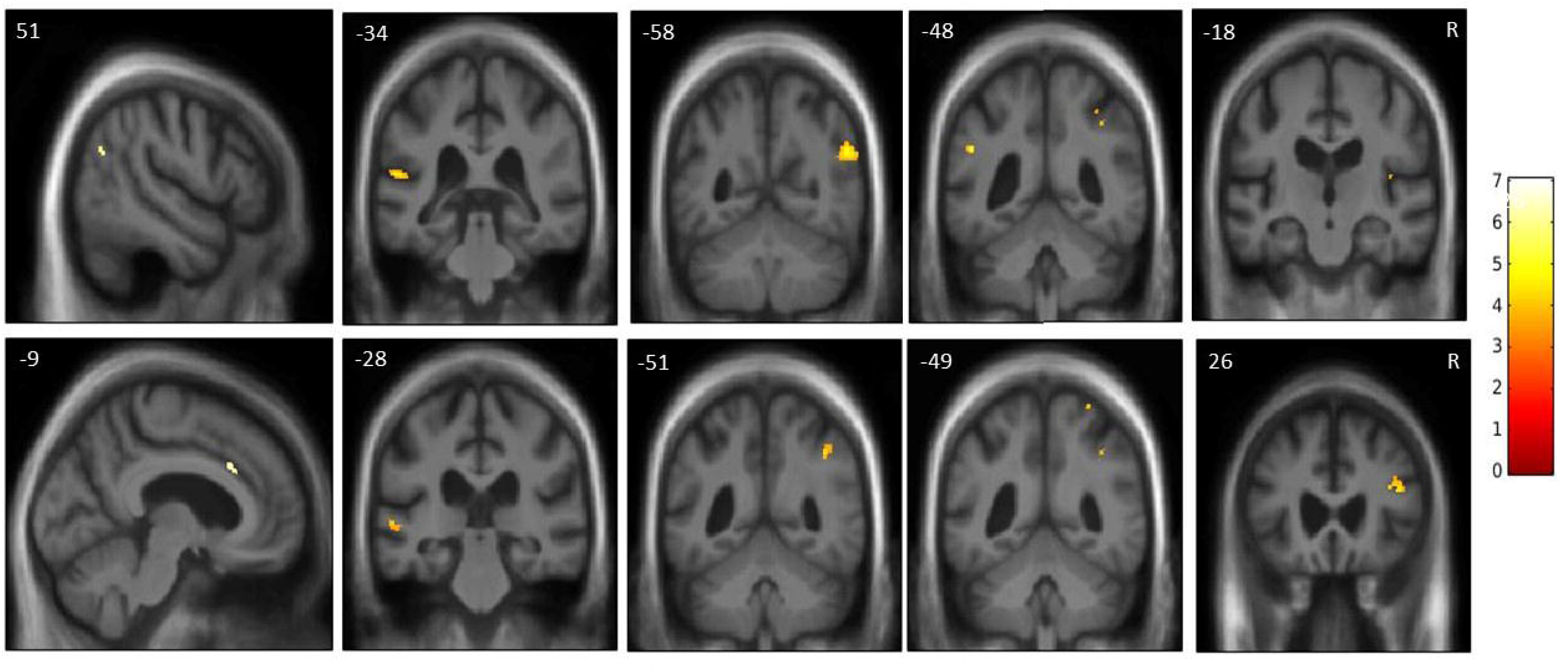
Statistical parametric maps of whole brain and regional grey matter atrophy associated with decreased dichotic listening performance in the combined patient cohort. Maps are rendered on sagittal sections of the group mean T1-weighted magnetic resonance image in MNI space. The first column includes sagittal scans of the significant regions thresholded at P < 0.05_FWE_ over the whole brain. The coronal scans show significant regions at P < 0.05_FWE_ (see also Table 4) within pre-specified neuroanatomical regions of interests (Figure S1), following an initial cluster-defining threshold (p<0.001). The colour bar (right) codes voxel-wise t-values. The plane of each section is indicated using the corresponding MNI coordinate (mm), and the right hemisphere is shown on the right side in the coronal sections.

Dichotic listening performance was significantly associated with regional grey matter volume in the left anterior cingulate gyrus and right temporo-parietal junction at whole-brain level (p_FWE_ <0.05), and with grey matter volume in the left planum temporale, right angular gyrus, left posterior supramarginal gyrus, right Heschl’s gyrus, left posterior superior temporal gyrus, right superior parietal lobule, and and right inferior frontal sulcus (all p_FWE_ <0.05 after correction for multiple voxel-wise comparisons within the relevant pre-specified neuroanatomical region of interest).

## Discussion

Here we have shown that dichotic listening is affected in patient groups with tAD and PPA syndromes relative to healthy individuals, and that these differences are significant after adjusting for peripheral hearing, auditory working memory (digit span forward), age and sex (Figure 1, Table 2). However, peripheral hearing function (better ear average, measured with pure-tone audiometry) was not significantly impaired in patients with tAD and PPA syndromes relative to healthy older individuals (Figure 1, Table 2), failing to replicate our previous findings of impaired peripheral hearing function in nfvPPA (Hardy et al., 2019). Furthermore, performance on the dichotic listening task in the combined patient group was positively and significantly correlated with MMSE and speech-based neuropsychological test scores, in comparison to non-significant correlations between pure-tone audiometry and neuropsychological test scores. These results suggest that dichotic listening performance indexes disease progression and active speech signal parsing, including neuropsychological functions that are dependent on this ability (e.g., auditory working memory). Our findings suggest that central hearing dysfunction (as measured with dichotic listening in this study) may index aspects of complex hearing that interact with other cognitive functions in the context of neurodegenerative syndromes. The findings in lvPPA corroborate previous evidence for impaired processing of speech sounds and auditory scene analysis in the atypical Alzheimer spectrum (Hardy et al., 2020; Johnson, Jiang, et al., 2020).

Significant neuroanatomical associations were observed between dichotic listening and grey matter volume in a cortical network that has previously been implicated in the processing of speech and disambiguation of competing sound signals (Table 3, Figure 3). Heschl’s gyrus has been shown to have a role in transforming acoustic signals for naturalistic speech processing (Khalighinejad et al., 2021). Left planum temporale and posterior superior temporal gyrus play critical roles in processing of speech (Jiang et al., 2023; Mesgarani et al., 2014; Möttönen et al., 2006), while right inferior frontal sulcus, angular gyrus and supramarginal gyrus have key roles in processing of speech in non-ideal listening conditions, selective attention to auditory features and domain-general resolution of competing sensory signals (Davis & Johnsrude, 2003; Golden et al., 2015; Jiang et al., 2021, 2023; Lemaitre et al., 2018; Szameitat et al., 2006). The right temporo-parietal junction has also been implicated in matching sensory inputs with internal representations, and attentional reorientation of unexpected stimuli (Bae et al., 2021). The right superior parietal lobules are functionally more typically associated with somatosensory and visuomotor processing (Gamberini et al., 2020; Wang et al., 2015), but have also previously been implicated in dichotic listening studies (Kompus et al., 2012), possibly reflecting their role in spatial processing and attentional allocation across sensory modalities (Vandenberghe et al., 2001; Wu et al., 2007). Left anterior cingulate works alongside the posterior cortical regions to decode spoken messages under challenging listening conditions and play a general role in allocating attentional resources (Gennari et al., 2018; Jiang et al., 2023; Obleser et al., 2007; Shenhav et al., 2013). Furthermore, these regions are all part of the core neural networks targeted in tAD and/or PPAs (Gorno-Tempini et al., 2011; Lombardi et al., 2021; Lorca-Puls et al., 2023).

**Table 3.** Neuroanatomical associations of dichotic listening performance in the combined patient cohort.

| Region | Peak (mm) |  |  | T score | P<sub>FWE</sub> |
| --- | --- | --- | --- | --- | --- |
|  | x | y | z |  |  |
| Left anterior cingulate gyrus | -9 | 24 | 33 | 6.58 | 0.004* |
| Right temporo-parietal junction | 51 | -63 | 27 | 6.00 | 0.018* |
| Left planum temporale | -54 | -34 | 15 | 4.85 | 0.004 |
| Right angular gyrus | 53 | -58 | 27 | 5.04 | 0.006 |
| Left posterior supramarginal gyrus | -50 | -48 | 28 | 4.59 | 0.017 |
| Right Heschl's gyrus | 41 | -18 | 12 | 3.65 | 0.023 |
| Left posterior superior temporal gyrus | -57 | -28 | 1 | 4.10 | 0.024 |
| Right superior parietal lobule | 38 | -48 | 42 | 4.03 | 0.032 |
| Right angular gyrus | 41 | -51 | 43 | 4.35 | 0.034 |
| Right superior parietal lobule | 30 | -49 | 70 | 3.97 | 0.037 |
| Right inferior frontal sulcus | 42 | 26 | 21 | 4.53 | 0.041 |

No associations were seen for peripheral hearing, corroborating previous findings of neuroanatomical correlates with central, but not peripheral hearing measures in patients with early neurodegenerative disease (Giroud et al., 2021). Importantly, previous studies reporting associations between pure-tone audiometry and neuroanatomical brain changes have typically been conducted in cognitively normal populations (Armstrong et al., 2019; Eckert et al., 2012; Lin et al., 2014; Parker et al., 2019; Ren et al., 2018; Rigters et al., 2017; Uchida et al., 2018), and possibly reflect down-stream effects of peripheral deafferentation on cortical reorganisation in otherwise healthy individuals. Our neuroanatomical analyses were restricted to patients, and it is possible that established neurodegenerative pathology attenuates any deafferentation-related associations.

Pure-tone audiometry (better ear average) was also not significantly associated with dichotic listening performance in the combined patient cohort or healthy individual group, suggesting that different mechanisms (i.e., peripheral and central, respectively) indeed underpin performance on these tasks. Furthermore, there were no significant correlations between better ear average and any neuropsychological variable. Here we failed to replicate previous reports of an association between peripheral hearing and cognitive performance (Loughrey et al., 2018) (Figure 2, Figure S2). One explanation is that we lacked power to detect these associations, but note that we did in fact see significant correlations between dichotic listening and each of the three tasks requiring speech perception for successful completion in the combined patient cohort (Figure 2, Figure S2). The associations reported in Loughrey et al. (2018)’s meta-analysis were weak, and seen for all-cause dementia but not Alzheimer’s disease, raising the possibility that the associations could be driven by intercurrent cerebrovascular disease. These results suggest that central brain hearing measures such as dichotic listening index aspects of complex hearing (Johnson, Marshall, et al., 2020) and highlight the importance of speech parsing mechanisms (beyond elementary sound detection) in driving cognitive test performance.

Healthy individuals and patients with lvPPA did show some evidence of a significant advantage for stimuli presented to the right ear (i.e., a right-ear advantage). However, our findings failed to replicate previous reports of significantly elevated dichotic right-ear advantage in patients with tAD, relative to healthy individuals of similar ages (Gates et al., 2008; Idrizbegovic et al., 2011; Tarawneh et al., 2022; Utoomprurkporn et al., 2020), and we failed to find evidence in support of our hypothesis that patients with lvPPA and nfvPPA would show an attenuated right-ear advantage relative to healthy controls. Possible explanations for these discrepancies include the relatively smaller sample sizes featured in the present study, and the fact that the paradigm employed here to detect right-ear advantage differed from ‘forced choice’ paradigms adopted in previous literature (i.e., asking participants to solely name numbers heard in right versus left ear) (Westerhausen & Kompus, 2018).

This study has limitations that suggest directions for future work. Here, we have considered just one measurement of peripheral and central hearing: future work should assess a wider range of peripheral and central hearing assays, including electrophysiology (Ferguson et al., 2023). The group sizes were relatively small, reflecting the rarity of the diseases in the case of the progressive aphasia participants. Future research should aim to corroborate the present findings in larger cohorts.

Clinically, this work has four main implications. Firstly, it highlights the importance of assessing central hearing alongside peripheral hearing in older adults at risk of dementia, and patients with established dementia: whilst our study did not assess this directly, it is possible that central hearing problems may underpin difficulties with hearing experienced in daily life, representing a form of ‘hidden’ hearing loss that would be missed with peripheral hearing assessments alone (Meyer et al., 2016; Slade et al., 2020). Relatedly, peripheral amplification with a hearing aid is not likely to adequately address central auditory processing deficits, and solutions customised for cognitively impaired populations (including those focusing on environmental modifications and communication partner training interventions) will be required (Kollmeier & Kiessling, 2018). Secondly, it adds to the growing evidence base that the primary progressive aphasias are associated with complex auditory phenotypes that go beyond language (Goll et al., 2010; Grube et al., 2016; Hardy, Buckley, et al., 2016; Hardy et al., 2017, 2018; Jiang et al., 2022, 2023; Johnson, Jiang, et al., 2020; Utianski et al., 2019). Thirdly, it speaks to the importance of isolating cognitive tasks requiring speech perception for successful performance from those that do not, lending support to approaches to adapt existing cognitive measures to overcome the associated confounds (Al-Yawer et al., 2019; Dawes et al., 2023). Finally, it consolidates previous work suggesting that central hearing tests may hold utility as early biomarkers for neurodegenerative diseases such as Alzheimer’s disease and primary progressive aphasia (Gates et al., 2011; Hardy, Marshall, et al., 2016; Jiang et al., 2023; Johnson, Marshall, et al., 2020; Stevenson et al., 2021). This points to an additional future need for clear care pathways from audiology to dementia diagnostic centres and care and support.

## Data Availability

The data that support the findings of this study are available on request from the corresponding author. The data are not publicly available because they contain information that could compromise
the privacy of research participants.

## Funding Acknowledgments

The Dementia Research Centre is supported by Alzheimer’s Research UK, Brain Research Trust, and The Wolfson Foundation. The work was supported by the Alzheimer’s Society (grant AS-PG-16-007 to J.D.W.), the Royal National Institute for Deaf People (G105 to J.D.W.), Alzheimer’s Research UK and the National Institute for Health Research University College London Hospitals Biomedical Research Centre. J.J. is supported by a Frontotemporal Dementia Research Studentship in Memory of David Blechner (funded through The National Brain Appeal). J.C.S.J. was supported by an Association of British Neurologists Clinical Research Training Fellowship. L.C. was supported by a UCL Research Excellence Scholarship. A.V. is supported by an NIHR Advanced Fellowship (NIHR302240). D.E.B. is supported by the Royal National Institute for Deaf People. C.R.M. is supported by a grant from Bart’s Charity and the National Institute for Health Research (NIHR204280). C.J.D.H. acknowledges funding from a RNID-Dunhill Medical Trust Pauline Ashley Fellowship (grant PA23_Hardy) and the National Institute for Health Research (NIHR204280). This study is funded by the NIHR [Invention for Innovation (NIHR204280)]. The views expressed are those of the authors and not necessarily those of the NIHR or the Department of Health and Social Care.

