## Supplementary for "Pure-tone audiometry and dichotic listening in primary progressive aphasia and Alzheimer’s disease"

**Correlations between dichotic listening and neuropsychological variables, accounting for peripheral hearing.** To ensure that significant associations between dichotic listening score and neuropsychological variables were not accounted for by peripheral hearing ability, we ran parallel partial correlation analyses adjusting for better ear average. In the combined patient cohort, dichotic listening total score was significantly correlated with arithmetic (r = 0.57, p < 0.001), forward digit span (r = 0.66, p < 0.001) and reverse digit span (r = 0.56, p < 0.001), after adjusting for better ear average.

**Table S1.** Peripheral listening characteristics of participant groups.

|  | **Healthy Individuals** | **AD** | **lvPPA** | **nfvPPA** | **svPPA** |
| --- | --- | --- | --- | --- | --- |
| **Peripheral hearing** | | | | | |
| Better ear average (BEA2, dB) (250Hz – 8000Hz) | 25.30 (10.56) | 32.46 (13.24) | 30.83 (16.78) | 32.65 (5.87) | 27.27 (9.03) |
| Better ear average (BEA3, dB) (4000Hz – 8000Hz) | 39.02 (19.48) | 46.45 (20.95) | 42.75 (22.62) | 49.32 (10.55) | 37.67 (14.86) |
| Better-hearing ear (E: L: R) (250Hz – 8000Hz) | 3:9:16 | 2:10:7 | 0:6:4 | 1:5:5 | 0:8:7 |
| Better-hearing ear (E: L: R) (4000Hz – 8000Hz) | 1:10:17 | 0:6:13 | 2:3:5 | 0:5:6 | 0:5:10 |

Mean (standard deviation) values and raw scores are presented (maximum possible value indicated in parentheses in test column). Unless otherwise indicated; significant differences from healthy individuals (p<0.05) are in **bold**. dB, decibel; E, equal; L, left; R, right.

**Table S2.** Statistical outputs for peripheral and dichotic listening models across participant groups.

|  | **Overall model** | **Age** | **Sex** | **Better ear average** | **Digit span forward score** | **Diagnosis** |
| --- | --- | --- | --- | --- | --- | --- |
| **Peripheral hearing** | | | | | | |
| Better ear average (dB) | **F(7,74) = 5.07, p<0.001** | **T=3.73, p<0.001** | T=1.79, p=0.078 | NA | T=1.25, p=0.216 | T=1.49, p=0.075 |
| Better ear average (dB) without digit span adjustment | **F(6,76) = 5.75, p<0.001** | **T=4.35, p<0.001** | T=1.73, p=0.087 | NA | NA | T=1.36, p=0.127 |
| Better-hearing ear (E: L: R) | Fisher’s exact = 0.547 | NA | NA | NA | NA | NA |
| **Dichotic listening** | | | | | | |
| Dichotic digits (total) | **F(8,73)=18.49, p<0.001** | T=0.63, p=0.340 | **T=2.65, p=0.010** | **T=2.63, p=0.010** | **T=5.16, p<0.001** | **T=2.23, p=0.001** |
| Dichotic digits (total) without digit span adjustment | **F(7,75) = 13.26, p<0.001** | T=1.77, p=0.081 | T=1.88, p=0.064 | T=1.66, p=0.102 | NA | **T=4.16, p<0.001** |
| Dichotic digits REA | F(8,73) = 1.15, p = 0.342 | NA | NA | NA | NA | NA |
| Dichotic digits REA without digit span adjustment | F(7,75) = 0.81, p = 0.580 | NA | NA | NA | NA | NA |
| Better-hearing ear (E: L: R) | Fisher’s exact = 0.589 | NA | NA | NA | NA | NA |

Statistical outputs of omnibus tests are presented, with significant values indicated in **bold**. To assess the impact of adjusting for forward digit span, models were run with and without this variable. There was a significant effect of diagnosis on dichotic listening total performance whether including this or not. After adjustment, there were significant differences between the healthy controls and each patient group, and the tAD and nfvPPA patient groups. Without adjustment, these significant differences remained, and the tAD group was significantly different to the lvPPA group (t=-2.01, p=0.048); the lvPPA group was significantly different to the svPPA group (t=2.64, p=0.010); and the nfvPPA group was significantly different to the svPPA group (t=4.46, p<0.001). dB, decibel; E, equal; L, left; lvPPA, logopenic variant primary progressive aphasia; NA, not applicable; nfvPPA, nonfluent/agrammatic variant primary progressive aphasia; R, right; REA, right ear advantage; svPPA, semantic variant primary progressive aphasia; tAD, typical Alzheimer’s disease.

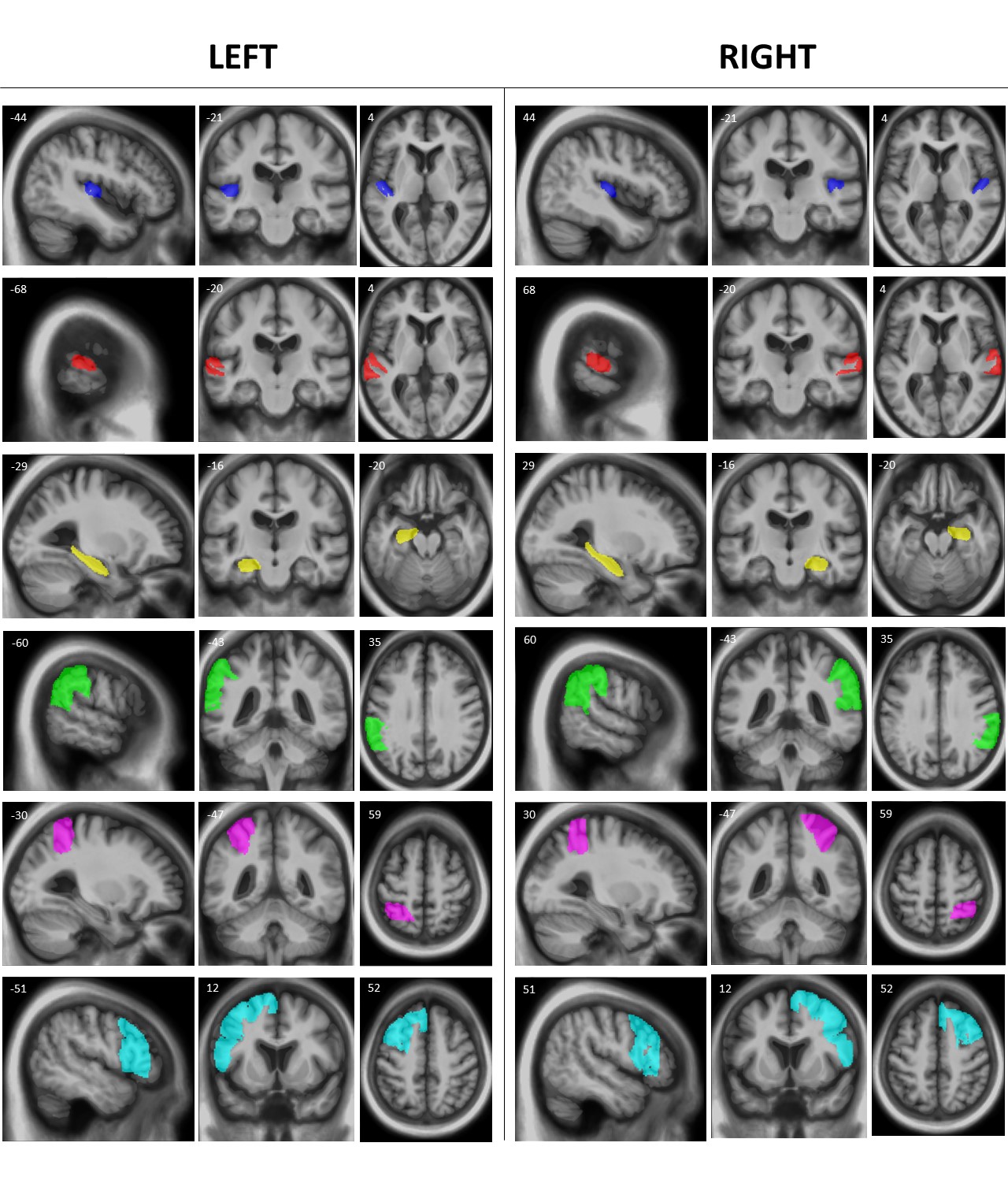

**Figure S1. Representative sections of neuroanatomical regions in both left and right cerebral hemisphere that were used for multiple voxel-wise comparisons correction in region-of-interest analyses (see text).** Regions are rendered on sagittal (left), coronal (middle) and axial (right) sections of the mean normalised brain template for the patient cohort; MNI coordinates of the plane of each section are shown. The neuroanatomical regions comprise Heschl’s gyrus (blue), the posterior superior temporal gyrus and planum temporale (red), medial temporal lobe (yellow), angular gyrus and supramarginal gyrus (green), superior parietal lobe (pink) and prefrontal cortex (cyan).

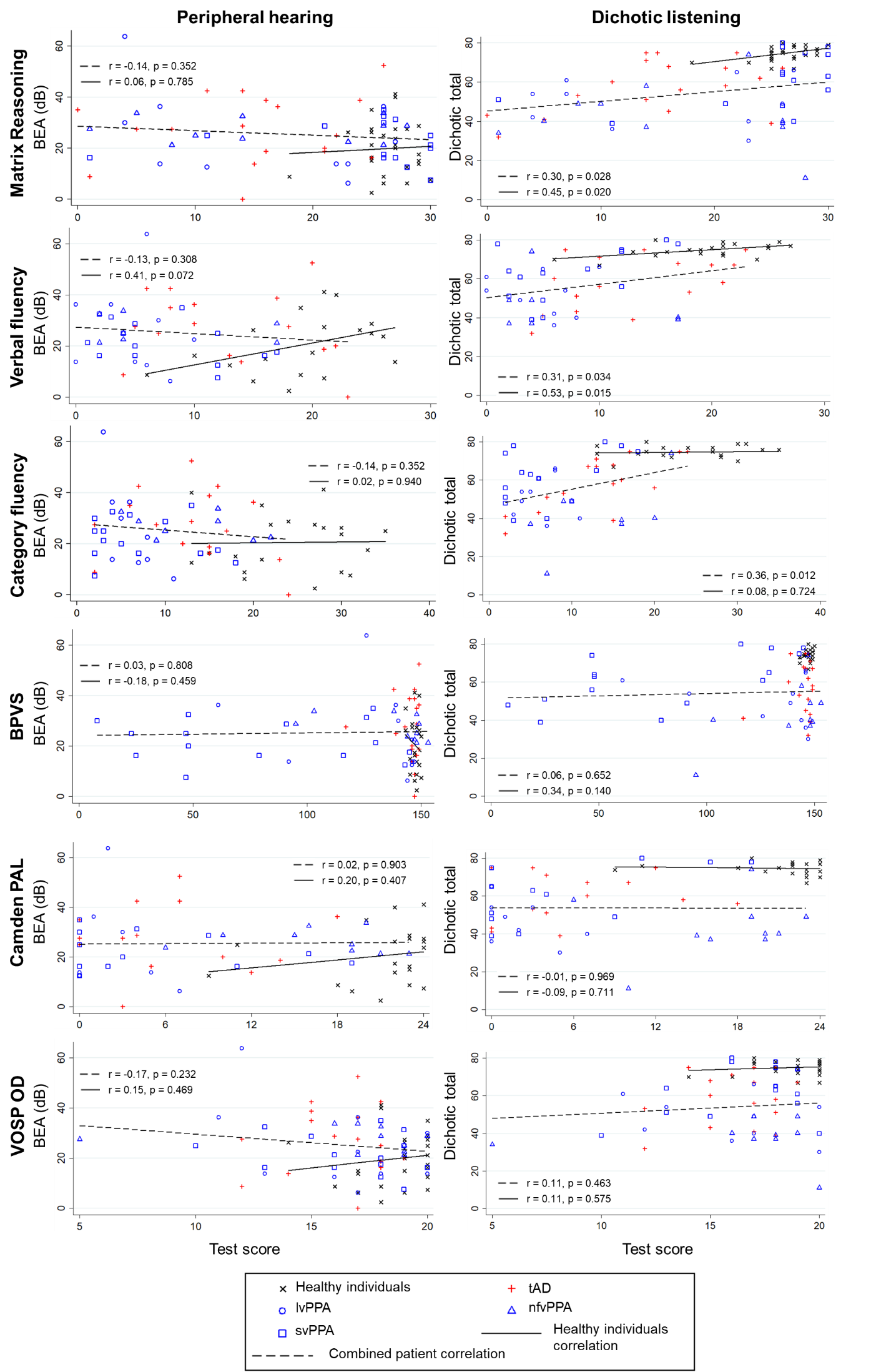

**Figure S2.** Scatter plots showing correlations between peripheral hearing ((better ear average); left panels) and dichotic listening (total score; right panels) with neuropsychological tests not requiring speech perception for successful performance in patient groups and healthy individuals. Diagnostic group membership is described in the key. For peripheral hearing, a lower pure tone audiometric average indicates better hearing so here negative *r* values would indicate that better peripheral hearing performance is associated with better psychometric performance. For central hearing, a higher dichotic digits total score indicates better hearing so here positive *r* values would indicate that better central hearing performance is associated with better psychometric performance. Correlations for tests requiring speech perception for successful performance are displayed in Figure 2. BEA, better ear average; control; healthy individual cohort; BPVS, British Picture Vocabulary Scale; tAD, typical Alzheimer’s disease; lvPPA, logopenic variant primary progressive aphasia; nfvPPA, nonfluent/agrammatic variant primary progressive aphasia; PAL, paired associates learning; svPPA, semantic variant primary progressive aphasia; VOSP OD, Visual Object and Space Perception Object Decision task.
